# Place of preperitoneal pelvic packing in severe pelvic traumatisms: About 20 cases performed in a French military level one trauma center

**DOI:** 10.1101/2021.02.09.21250850

**Authors:** Hardy Julie, Coisy Marie, Monchal Tristan, Bourguoin Stéphane, Long Depaquit Thibaut, Chiron Paul, Mickael Cardinale, Hornez Emmanuel, Balandraud Paul, Savoie Pierre-Henri

## Abstract

**Background:** The overall mortality of hemodynamically unstable pelvic fractures is high. Hemorrhage triggers off the Moore lethal triad. Hemostatic management during the golden hour is essential. Combined with pelvic stabilisation, preperitoneal pelvic packing (PPP) is proposed to control venous and bony bleeding, while arterioembolisation can stop arterial bleeding. No international consensus has yet prioritized these procedures. The aim of this study was to analyse a serie of PPP in a military level one trauma center and propose an algorithm for hemodynamically unstable pelvic traumas regardless of the military facility.

**Method:** From January 2010 to December 2020, every patient from our military institution with a hemodynamically unstable pelvic fracture underwent PPP combined with pelvic stabilisation. Before 2012 data were retrospectively collected from database (PMSI), after 2012 data were prospectively recorded in our polytrauma database and retrospectively analysed. The care algorithm applied focused on hemodynamic status of polytraumatised patients on admission. Primary criteria were early hemorrhage-induced mortality (<24h) and overall mortality (<30d). Secondary criteria were systolic blood pressure (SBP) and red blood cells (RBC) units administered.

**Results:** 20 patients with a pelvic fracture had a PPP. Mean age was 49,65 +/-23,97 years and median ISS was 49 (31; 67). The decrease of blood transfusion and increase of SBP between pre- and postoperative values were statistically significant. Eight patients (40%) had postoperative arterial pelvic blush and 7 patients were embolised. The early mortality by refractory hemorrhagic shock was 25% (5/20). Overall mortality at 30 days was 50% (10/20).

**Conclusion:** PPP is a quick, easy, efficient and safe procedure. It can control venous, bony and sometimes arterial bleeding. PPP is part of damage control surgery and we propose it in first line. Angio-embolization remains complementary. Besides, PPP is the only means available in precarious conditions of practice, notably in military forward units.

## INTRODUCTION

Traumatic pelvic ring fractures with hemodynamic instability are life-threatening injuries. The overall mortality for pelvic traumatisms with shock is 40-90% despite multidisciplinary approaches (1,2). Hemorrhage is mainly caused by low-pressure venous plexi and bones, then by high-pressure arterial lesions (3). Because of hemorrhage, patients quickly enter the Moore lethal triad of acidosis – hypothermia – coagulopathy. Early hemostatic management during the golden hour is essential. There is no way to diagnose the precise source of pelvic bleeding responsible for hemodynamic instability. Since the 80’s, angio-embolization (AE) has become a standard however it stops only arterial bleeding. In more recent years, pelvic packing combined with pelvic stabilization has been proposed to control venous and bony bleeding. Techniques and indications of pelvic packing have evolved, and the current technique is an early and exclusive retroperitoneal approach combined with external fixation of the pelvis, as described first by European trauma teams in the 2000’s (3, 4) and then by American trauma teams (5).

The aim of this study was to review our series of preperitoneal pelvic packing (PPP), assess the efficiency of this approach, compare it to literature and propose an algorithm for the management of hemodynamically unstable pelvic fractures.

## MATERIALS AND METHODS

### 1. Study design

This is a monocentric retrospective observational study. All patients who underwent PPP for hemodynamic instability on traumatic pelvic fracture were included and analyzed. From January 2010 to December 2011, data of the cohort were retrospectively collected from a PMSI research. Since January 2012, data were prospectively collected in a trauma base to December 2020 in a French level I trauma reference center (Sainte-Anne Military Teaching Hospital (SAMTH), Toulon, France).

### 2. Management

#### 2.1 Initial management

Prehospital assessment was carried out using the Vittel criteria (6)(Table 1). Resuscitation was managed according to the national guidelines edited by SFAR (société française d’anesthésie réanimation). A pelvic binder was set up when pelvic disruption was suspected.

**Table 1:** Vittel criteria. A patient with <u>one</u> of the listed criteria was considered as a severe trauma and was transferred to a trauma center.

| Steps | Severity criteria |
| --- | --- |
| Vital signs | Glasgow coma scale < 13<br>Systolic blood pressure < 90 mmHg<br>Saturation O<sub>2</sub> < 90% |
| Evidence of high-energy trauma | Ejection from automobile<br>Death in same passenger compartment<br>Falls > 6 m<br>Victim thrown or crushed<br>Global assessment of the trauma (aspect of the crashed vehicle, vehicle telemetry data consistent with high risk of injury, no motorcycle helmet, no seat belt)<br>Blast |
| Anatomy of injury | Penetrating trauma of head, neck thorax, abdomen, pelvis, thigh, and arm<br>Flail chest<br>Severe burns, smoke inhalation<br>Pelvic bone fracture<br>Suspicion of medullar trauma<br>Amputation proximal to wrist or ankle<br>Acute ischemia of the limb |
| Pre-hospital resuscitation | Intubated and mechanically ventilated patients<br>Intravenous fluids > 1000 mL (colloids)<br>Catecholamine<br>Anti-shock trousers inflated |
| Special patient or system considerations | Age > 65 yr<br>Heart failure<br>Respiratory failure<br>Pregnancy > 12 weeks |

The algorithm applied in our level I trauma center focused on the hemodynamic status of severely injured patients at admission (Figure 1). Hemodynamic instability was defined as systolic blood pressure < 90mmHg despite fluid therapy (> 2L crystalloids) and transfusion of 2 red blood cells (RBC). As in all French military hospitals, plasma was available as fresh frozen plasma (FFP) and lyophilized plasma (FLyoP).

**Figure 1.**
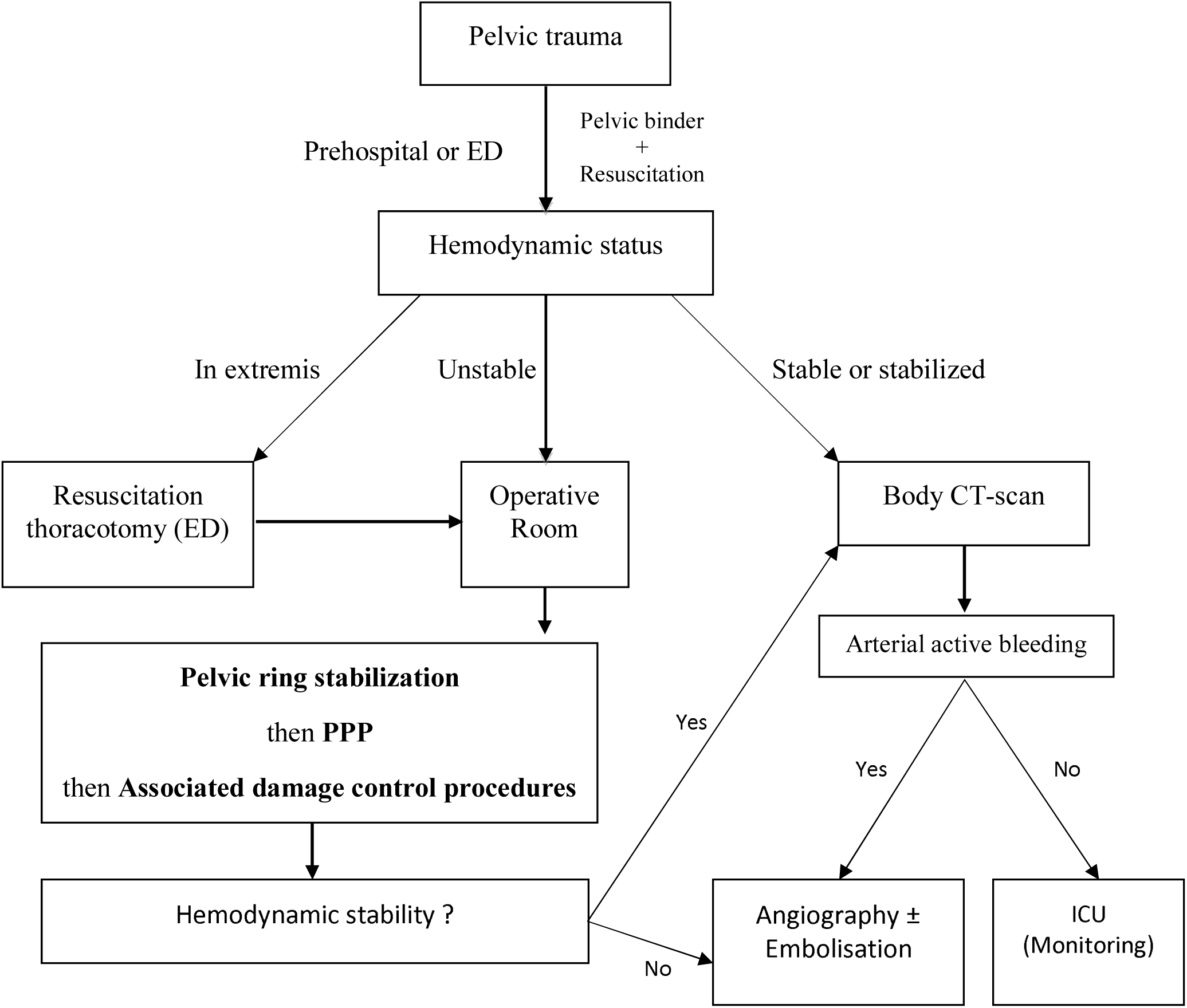
Algorithm in Sainte Anne Military Hospital (level 1 trauma center) for polytraumatized patients with pelvic fracture (ED = Emergency Department; ICU = Intensive Care Unit)

Patients who had cardiopulmonary arrest in the emergency department (ED) were considered “in extremis” and underwent resuscitation thoracotomy with aortic cross-clamping before transfer to the operating room (OR). For other patients, chest and pelvic plain X-ray films and focused abdominal sonography for trauma (FAST) were performed during resuscitation management.

At this stage, hemodynamically stable or stabilized patients had a complete lesional assessment by body computed tomography (CT) before being given any treatment.

Unstable patients with identified pelvic disruption were transferred to the OR to undergo PPP and orthopedic damage control surgery. From 2013, the algorithm was partially modified by a systematic use of surgical stabilisation (C-clamp or external fixation).

Injuries were coded according to the abbreviated injury scale for the calculation of injury severity score (ISS). Tile classification was used to categorize fracture patterns (7). A severely injured patient was defined as injured to 2 or more organ systems and ISS > 15. An “in extremis” patient was defined as a patient under cardiac arrest in the ED.

#### 2.2 Surgical technique of PPP (Figure 2)

A 6 to 8 cm midline incision was made from the pubis in direction of the umbilicus. Skin, subcutaneous tissue and the midline fascia were opened. The retroperitoneum plane was easy to find thanks to its expansion by hematoma and clots. The peritoneum was left intact. The bladder was retracted, away from the pubis. To have efficient packing, three or more large radiopaque laparotomy pads were placed in each side in the space between the pelvic girdle and the peritoneum, from the sacroiliac joint onto the paravesical and retropubic zone. The pads were placed directly near the branches of the internal iliac artery and the pelvic venous plexi, lateral to the sacrum. No direct ligation of the hypogastric artery was performed. A cystostomy was placed after packing. The outer fascia was closed with a running suture and the skin incision was stapled. If necessary, abdominal damage control surgery was performed by a subsequent and separated abbreviated laparotomy.

**Figure 2.**
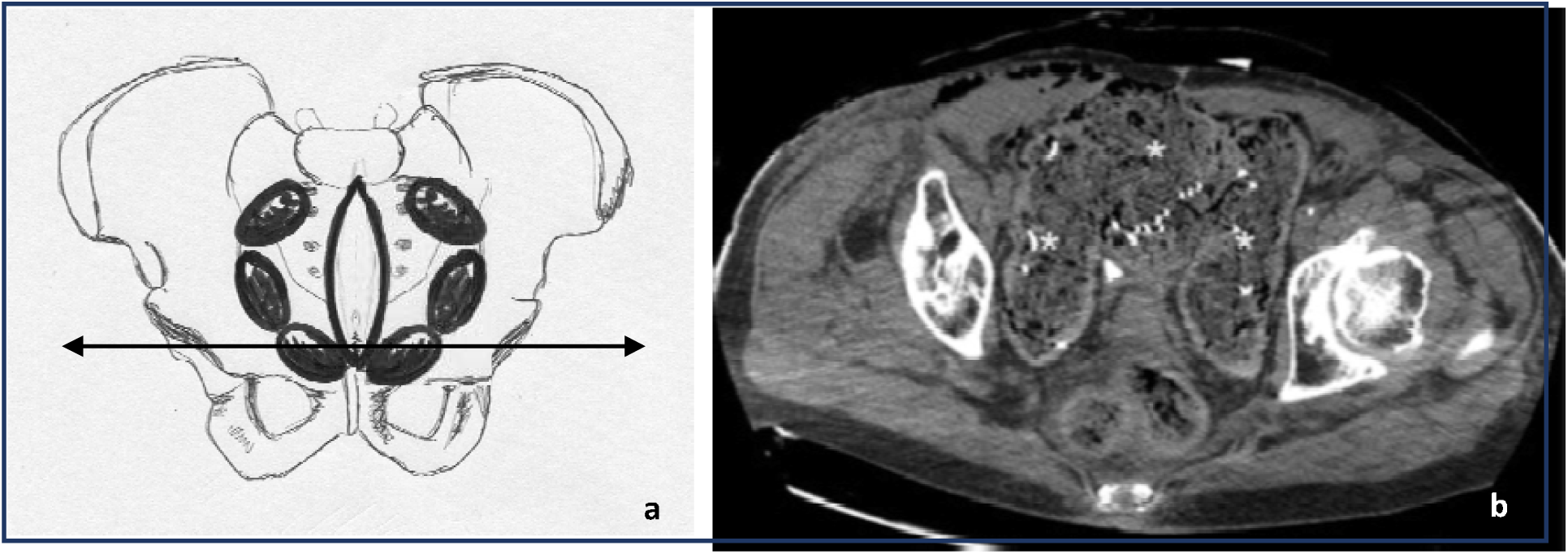
Preperitoneal pelvic packing : Technical aspects. Diagram showing the disposition of the incision and pads (a) and corresponding pelvic-CTscan slice (b) performed after preperitoneal pelvic packing : The laparotomy gauze (*) fill the lateral rectal fossae, the space of Retzius and the retro-inguinal space of Bogros, compressing the venous plexuses against the bony pelvis.

#### 2.3 Additional Hemostatic Procedures

Angiography was subsequently performed in case of persistent bleeding or persistent hemodynamic instability after surgical procedure. If needed, selective AE was performed.

### 3. Data collection and statistical analysis

From January 2010 to December 2011 we searched patients via PMSI (programme de médicalisation des systèmes d’informations) codes crossing all types of pelvic fractures with hemodynamic instability. From January 2012, all patients severely injured were prospectively registered in our institution’s pelvic trauma registry (authorized by the French National Commission on Data Protection (CNIL) listed under the reference MR-001 No. 1578624vO and in Traumabase® (www.traumabase.eu). Traumabase.eu, created in 2012, is a French network database dedicated to all admissions due to whatever trauma. Traumabase® obtained approval from the Advisory Committee for Information Processing in Health Research (CCTIRS, 11.305bis) and from the CNIL (authorization 911461) and meets the requirements of the local and national ethics committees (Comité de Protection des Personnes, Paris VI). According to the French law (8), our study was approved by the local SAMTH ethics commitee (IRB 0011873-2020-02) by decision on 04/14/2020.

We identified among them those who had pelvic fracture and those who underwent PPP. Patient information -such as age, sex, injury mechanism, ISS, associated injuries, time of emergency resuscitation, time to OR, time for completion of PPP, mean stay in intensive care unit and hospital stay -was extracted from the database. Other specific data were retrospectively reviewed on electronic and paper records: ED physiological parameters (systolic blood pressure (SBP)), heart rate, hemoglobin rate, lactates, pH, platelets, prothrombin time, temperature), type of pelvic fracture, SBP before and immediately after PPP, amount and time of RBC transfusion, associated surgical procedures, AE, complications, time and cause of death. Surgical duration was considered for PPP alone.

Primary criteria were early hemorrhage-induced mortality (<24h) and overall mid-term mortality (<30d). Secondary criteria were SBP and RBC units administered. A sub-group analysis compared the characteristics of patients who died of hemorrhagic shock to the ones who survived 24 hours after traumatism.

All statistical analyses were conducted using GraphPad Prism 8.0 (GraphPad Software Inc, San Diego, California). Categorical variables were presented as numbers and percentages, and continuous variables were presented as median and interquartile range (IQR) or mean ± standard deviation (SD). Univariate analyses were performed using the Pearson’s chi-squared test or Fisher’s exact test for categorical variables, and Mann Whitney U test for continuous variables, as appropriate. A two-sided p-value of less than. 05 was considered to indicate statistical significance.

## RESULTS

### 1. Population characteristics

From 2010 January to 2020 December, 287 severely injured patients were admitted and had pelvic fracture (table 2). Among them, 20 patients underwent PPP because of associated hemodynamic instability. In this group, the object of our study, 9 patients were initially hemodynamically unstable, 7 became unstable after the body CT, and 4 were unstable after their transfer from peripheral hospitals. These 4 patients were transferred to be angio-embolized for a blush identified on the initial body CT or to be taken care of for injuries that were not technically possible in the peripheral hospital. The peripheral hospitals were 85 km for Draguignan and 50 km for Brignoles. Heliport transport was used.

**Table 2.**
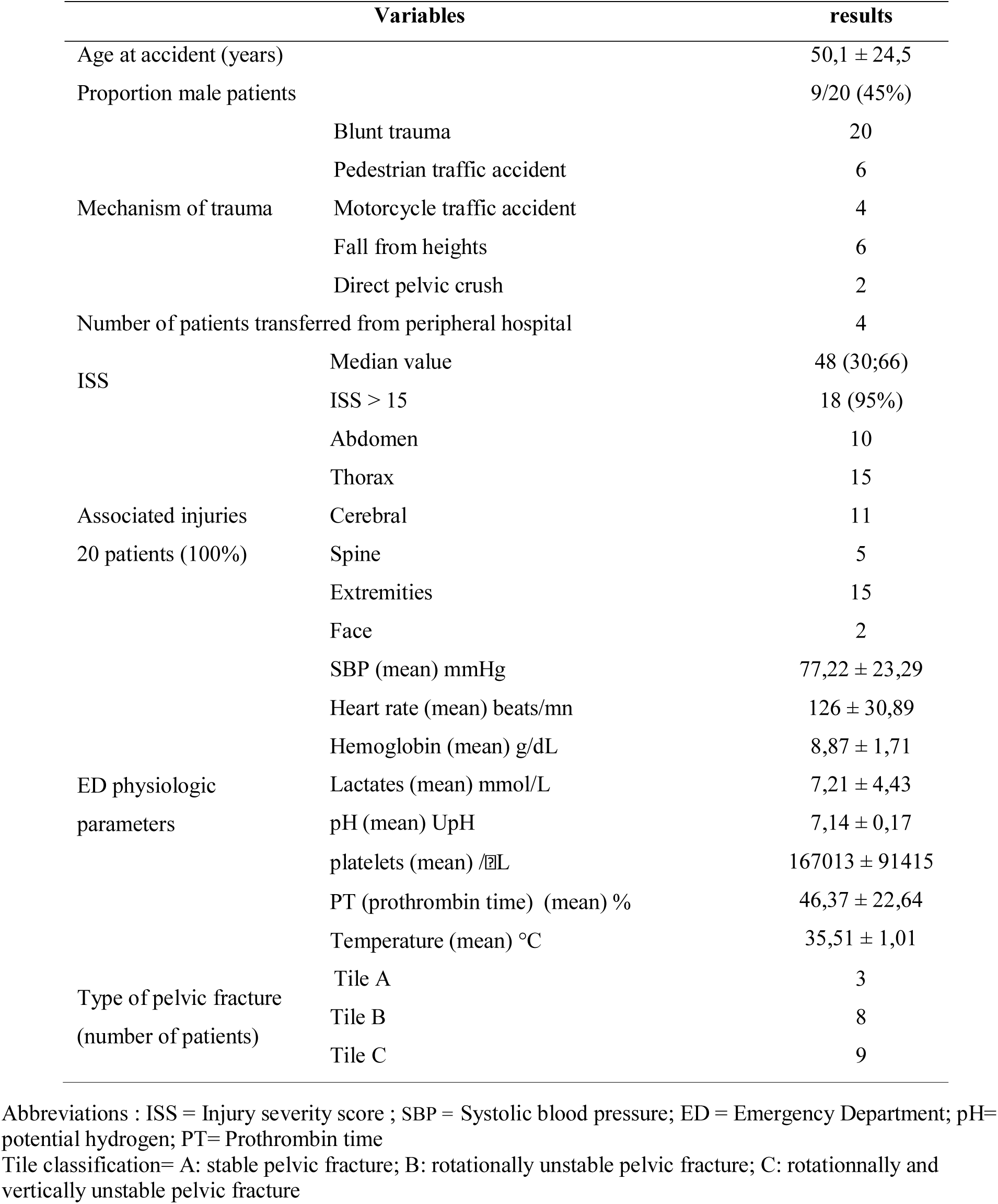
Population characteristics (n=20)

### 2. ED physiologic parameters and resuscitation

All physiologic parameters confirmed the severity of the hemorrhagic shock of the patients in this study, in particular acidosis with a mean pH 7.15 and mean lactacidemia 7,29 (table 2). The ratio RBC / plasma was 2/1. Tranexamic acid and fibrinogen were systematically administered.

### 3. Hemorrhage-control interventions (Table 3)

Indication for PPP was immediate for 13 patients with hemodynamic instability (10 directly admitted and 3 transferred patients who became secondary unstable), and for 7 patients, PPP was secondary after completion of CT. Considering the 13 at once hemodynamically unstable patients, PPP was completed within 36,25+/-31,56 min from ED admission. Concerning secondary hemodynamic patients, PPP was completed within 109,29+/-39,42 min. The mean duration for completion of PPP from incision to skin closure was 23,3+/-10,88 min. We found a statistically significant difference between preoperative SBP (78,24 mmHg) and postoperative SBP (118,82 mmHg) (p<0.001). The median transfusion rate decreased significantly from 4 RBCs administered preoperatively to 2 RBCs transfused within the 24h following PPP (p=0.0231). After PPP, 7 patients underwent angiography followed by embolization: 2 for a preoperatively documented blush at CT-scan, 4 for a blush diagnosed on the postoperative whole body CT-scan (distal hepatic artery, segment 4 of the liver, hypogastric artery in two patients) and 1 for persistent hemodynamic instability. They underwent depacking at 48 hours and only 1 needed a repacking because of a coagulopathy.

**Table 3.**
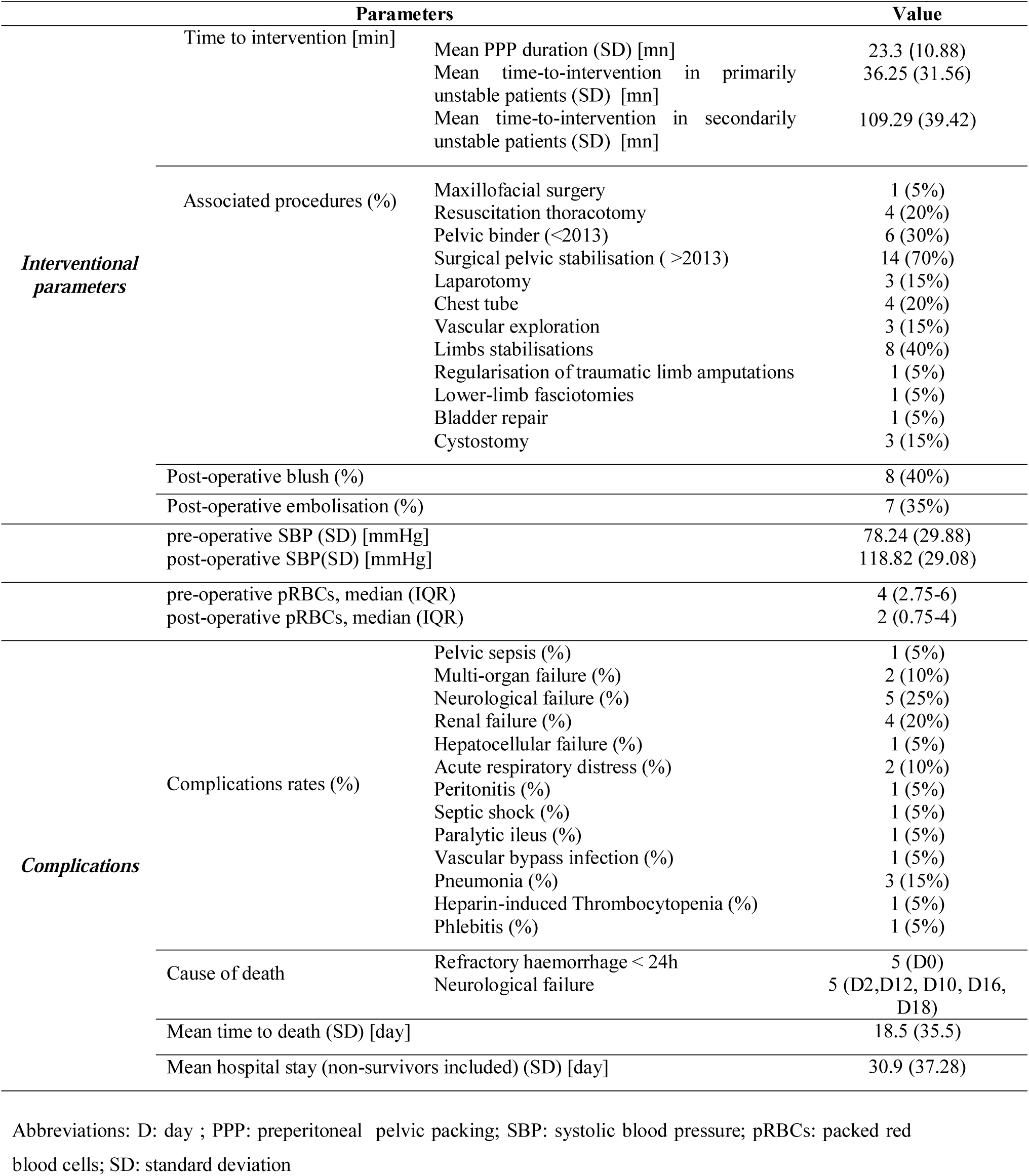
Peri-operative parameters and complications. 8 extremities stabilizations : 3 femoral external fixators, 2 tibial external fixator, 3 lower limb tractions 3 vascular explorations : 1 scarpa injury, 1 ilio-femoral bypass, 1 dissection segment 3 aorta 1 peritonitis : undetected traumatic Small bowel injury 1 septic shock : mesenteric ischemia

### 4. Mortality

Overall mortality was 50% (10/20). Five patients (25%) died from exsanguination within 24h hours. Among them, one had several cardiac arrests in the ED, and 4 underwent resuscitation thoracotomy. Five patients (25%) died within 30 postoperative days, all for neurologic failure (table 3).

### 5. Related morbidity

We identified 1 pelvic sepsis, which was not lethal. Other complications were non-specific (renal failure : 4) or related to trauma (neurologic failure : 5). Postoperative outcomes are shown in Table 3.

### 6. Sub-group analysis: early mortality (Table 4)

Prothrombin time (PT) was significantly lower (p=0.047) and age was significantly higher (p=0.032) in the patients who died of hemorrhagic shock within the first 24 hours. Preoperative blood transfusion (p=0.072), postoperative blood transfusion (p=0.813), postoperative SBP (p=0.860) and type of fracture (p=0.929) were not statistically different (Table 4).

**Table 4.**
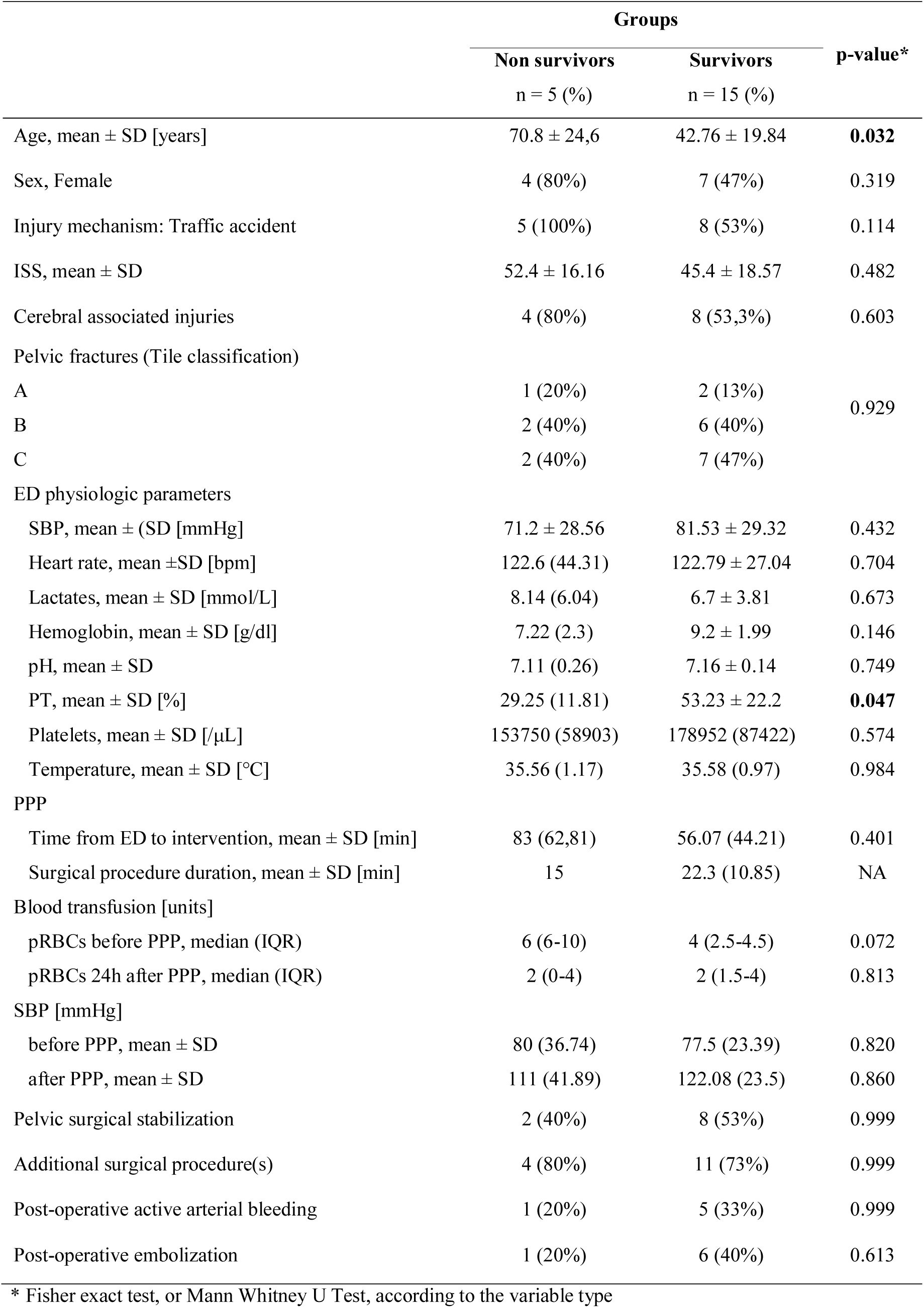
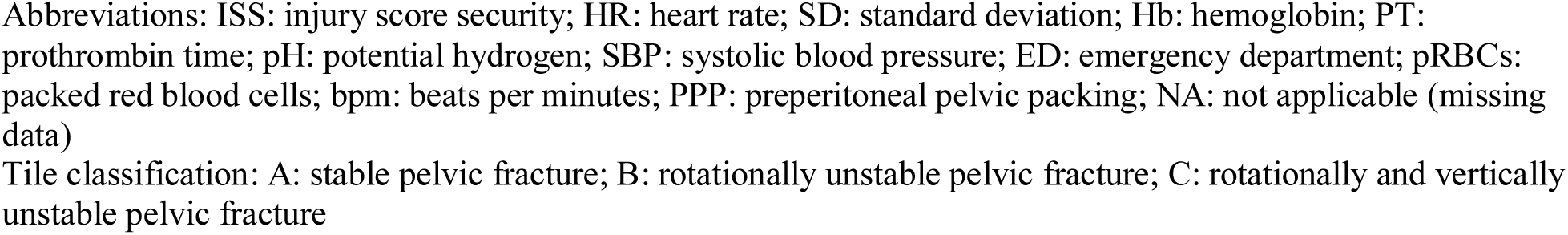
Comparison between survivors and non-survivors at h24.

|  | Groups |  | p-value* |
| --- | --- | --- | --- |
|  | Non survivors<br>n = 5 (%) | Survivors<br>n = 15 (%) |  |
| Age, mean ± SD [years] | 70.8 ± 24,6 | 42.76 ± 19.84 | **0.032** |
| Sex, Female | 4 (80%) | 7 (47%) | 0.319 |
| Injury mechanism: Traffic accident | 5 (100%) | 8 (53%) | 0.114 |
| ISS, mean ± SD | 52.4 ± 16.16 | 45.4 ± 18.57 | 0.482 |
| Cerebral associated injuries | 4 (80%) | 8 (53,3%) | 0.603 |
| Pelvic fractures (Tile classification) |  |  |  |
| A | 1 (20%) | 2 (13%) | 0.929 |
| B | 2 (40%) | 6 (40%) |  |
| C | 2 (40%) | 7 (47%) |  |
| ED physiologic parameters |  |  |  |
| SBP, mean ± (SD [mmHg] | 71.2 ± 28.56 | 81.53 ± 29.32 | 0.432 |
| Heart rate, mean ±SD [bpm] | 122.6 (44.31) | 122.79 ± 27.04 | 0.704 |
| Lactates, mean ± SD [mmol/L] | 8.14 (6.04) | 6.7 ± 3.81 | 0.673 |
| Hemoglobin, mean ± SD [g/dl] | 7.22 (2.3) | 9.2 ± 1.99 | 0.146 |
| pH, mean ± SD | 7.11 (0.26) | 7.16 ± 0.14 | 0.749 |
| PT, mean ± SD [%] | 29.25 (11.81) | 53.23 ± 22.2 | **0.047** |
| Platelets, mean ± SD [μL] | 153750 (58903) | 178952 (87422) | 0.574 |
| Temperature, mean ± SD [$^{\circ}$C] | 35.56 (1.17) | 35.58 (0.97) | 0.984 |
| PPP |  |  |  |
| Time from ED to intervention, mean ± SD [min] | 83 (62,81) | 56.07 (44.21) | 0.401 |
| Surgical procedure duration, mean ± SD [min] | 15 | 22.3 (10.85) | NA |
| Blood transfusion [units] |  |  |  |
| pRBCs before PPP, median (IQR) | 6 (6-10) | 4 (2.5-4.5) | 0.072 |
| pRBCs 24h after PPP, median (IQR) | 2 (0-4) | 2 (1.5-4) | 0.813 |
| SBP [mmHg] |  |  |  |
| before PPP, mean ± SD | 80 (36.74) | 77.5 (23.39) | 0.820 |
| after PPP, mean ± SD | 111 (41.89) | 122.08 (23.5) | 0.860 |
| Pelvic surgical stabilization | 2 (40%) | 8 (53%) | 0.999 |
| Additional surgical procedure(s) | 4 (80%) | 11 (73%) | 0.999 |
| Post-operative active arterial bleeding | 1 (20%) | 5 (33%) | 0.999 |
| Post-operative embolization | 1 (20%) | 6 (40%) | 0.613 |

\* Fisher exact test, or Mann Whitney U Test, according to the variable type
Abbreviations: ISS: injury score security; HR: heart rate; SD: standard deviation; Hb: hemoglobin; PT: prothrombin time; pH: potential hydrogen; SBP: systolic blood pressure; ED: emergency department; pRBCs: packed red blood cells; bpm: beats per minutes; PPP: preperitoneal pelvic packing; NA: not applicable (missing data)
Tile classification: A: stable pelvic fracture; B: rotationally unstable pelvic fracture; C: rotationally and vertically unstable pelvic fracture

## DISCUSSION

Overall mortality of pelvic fracture with hemodynamic instability remains high despite a multidisciplinary approach: from 40 to 90-100% for the most injured patients (1, 2, 9, 10). The challenge is to stop the bleeding as soon as possible (11). Advantages of PPP in the first line is defended by a growing number of traumatologic teams following results of recent series (12-14). Several reasons explain why our institution has adopted this attitude.

Mechanical impact of PPP on hemorrhage should be considered. The specificity of pelvic trauma is the multiple sources of bleeding (venous plexus, bones and arteries). Like in hepatic trauma, PPP is efficient on low-pressure injuries and AE on arterial injuries. Majority of anatomical and radiological studies conclude that the main component of pelvic hemorrhage comes from venous and cancellous bone surfaces from fractures and sacro-iliac joint disruptions (15). Conversely a few studies, mainly radiological, highlight the arterial component (16). We consider that in pelvic fractures, there is always a venous and a bone component in bleeding. So in our institution, PPP is performed as soon as possible, sometimes before bodyCT from the resuscitation room of the ED. Some authors consider PPP as invasive mainly because of septic risks. In this series, infectious complications attributed to PPP were limited (5%) and never fatal. The pelvic binder is left until the OR to provide temporary bone stability and open book effect. The type of fracture does not predict the amount of blood loss and therefore it is not a parameter included in our algorithm. It has been demonstrated that pelvic stabilization decreases pelvic volume by 10 to 20% (15). It is moreover a quick procedure, lasting 15 minutes (3). In OR, the pelvic strapping is relaxed and a surgical pelvic stabilization completes the damage control surgery.

Physiological impact of PPP is also an important parameter to analyze. Blood transfusion has been proved an important mortality risk factor because favoring multi-organ failure (MOF) (5). Reducing the need for transfusion is a compelling objective. In this series, PPP has decreased significantly the amount of transfused packed RBC between pre- and postoperative (p=0.0231) and only 1 MOF occurred, as observed in other series (9). Likewise, the impact of PPP on SBP was evident, as shown in other series where a statistical difference was found between pre- and postoperative SBP; they all demonstrated a statistically significant increase of SBP (12-14), as we did (p<0.001). A recent study (17) suggests that kaolin-impregnated hemostatic gauze in PPP could reduce the need for additional packed RBC transfusions in patients with hemodynamic instability due to severe pelvic fractures. This could be a useful improvement to add to reduce the transfusions.

As regards to technical aspects, our surgical procedure evolved over its 10 years duration, due to improvement on the algorithm. For example, pelvic stabilization was modified with the purchase of a C-clamp in 2013, excluding the simple pelvic contention by binder. About urologic management, the first patients of this study had a trans-urethral catheter to drain the bladder. We know that it can increase potential urethral injuries and that bladder distension contribute to pack the retroperitoneal hematoma. That is why we decided to place a cystostomy at the end of the surgical procedure. Besides, in an anatomical study, Grimm et al. showed that a laparotomy significantly decreased retroperitoneal pressure (18). Hence our decision to perform a short separated incision to leave the peritoneum intact and to avoid any communication between the intraperitoneal and the retroperitoneal spaces. Since a few months, a new procedure by REBOA (Resuscitative Endovascular Balloon of the Aorta) set up for “in extremis” patients but we have not used it yet in unstable pelvic fractures. Nevertheless REBOA is efficient on arterial bleeding but not in venous or bone bleeding. Therefore it could be complementary with PPP and not competitive.

The EAST guidelines (Eastern Association for the Surgery of Trauma) (19) recommend primary AE, whereas the European ones recommend pelvic stabilization and PPP first (20, 21). However, patients undergoing a diagnostic angiography do not all have active arterial bleeding and few of them need embolization (9). Besides, its efficiency is recognized on angiographic criteria and not on blood pressure stabilization criterion (22). Neither has angiography an impact on the global amount of RBC transfusion (9). Finally, this procedure is time-consuming and delays resuscitation, even in high volume centers when AE is available. It is reasonable to propose it when embolization probability is high. Osborn et al. also found only 50% of active arterial pelvic bleeding in their ANGIO group, which means that 50% of their primary angiographies were non-therapeutic. Furthermore, mortality after primary angiography remains high: from 26% up to 69% (13). For all these reasons, its place in first intention care is controversial. The European guidelines also suggest systematic postoperative angiography. In the six major PPP series published (4, 13, 14, 23), postoperative angiography was performed only if there were signs of further bleeding: persistent hemodynamic instability, persistent need for RBC transfusion or persistent high level of lactates. In our series, postoperative angiography was not systematic. Most patients had postoperative vascular cartography by whole body CT when hemodynamically stable. Angiography was performed for patients with signs of persistent bleeding or who had a blush on their body CT. Almost these guidelines, an american referent trauma center validated recently PPP for reducing mortality in patients with life-threatening hemorrhage due to unstable pelvic fractures (24).

Nowadays, with the golden hour principle, the management of hemorrhagic patients is guided by the will to shorten the delay for these patients to reach hemostatic procedures. Indeed, it has been shown that a prolonged time before achieving hemostasis has an adverse effect on patients’ survival (4) Time to reach the interventional room (IR) for angiography has been described from 102 min to 130 min (13) and the procedure duration must be added, from 84 min (13) to 5.5 h (22). On the contrary, performing a PPP is fast : time from ED to OR is limited (36.25 min in this study, 41 to 78min for other authors (4, 12, 13) and surgical procedure is short (23.3 min in this study). Osborn et al. demonstrated a statistically significant difference (p=0.041) of arterial bleeding between their ANGIO group (50%) and PACK group (15%) (13). This suggested that PPP may have helped in controlling abundant pelvic arterial hemorrhage and selecting patients who could most benefit from pelvic angioembolization. Furthermore, it makes choosing between OR and IR irrelevant. Indeed, IR is not a suitable resuscitation environment for unstable trauma patients. In OR, additional surgical damage control procedures may be performed simultaneously. This is particularly beneficial, since up to 90% of patients with unstable pelvic fractures have associated injuries and 50% of patients have other sources of major hemorrhage than pelvic fractures (22).

Finally, in rural resource-poor settings, interventional radiology is not always available for selective angiography. In these cases, PPP combined with pelvic stabilisation should be performed for a patient with hemodynamic instability before his transfer to another facility. Therefore, every civilian surgeon should know this technique. The vast majority of military deployed medical facilities are also resource constrained. Considering the technical platform in a French forward surgical unit, PPP is the only appropriate procedure praticable whatever the battlefield pelvic injuries. Denver’s trauma surgical team, indeed, concluded recently PPP was effective for hemorrhage control in patients with open pelvic fractures, regardless mechanisms penetrating or blunt (25). French military surgeons propose PPP in combination with percutaneous external fixation for controlling life-threatening haemorrhage in unstable patients with pelvic fractures penetrating or not. They applied these guidelines in the first line of management in their level one trauma centers and like exclusive treatment in the austere environment of a french forward unit. This procedure should be considered early in patients with military pelvic trauma and major haemorrhage, as part of damage control surgery in war context.

## CONCLUSION

This study is the first PPP series from a French trauma center. It confirms that PPP is a quick, easy, efficient and safe procedure. Its efficiency on venous and bony bleeding has been proven. Even if there is an arterial component of bleeding, PPP contributes to stop bleeding and sometimes avoid arteriography. It is part of damage control surgery and we propose it in the first line. AE remains complementary, and is to be done second. Besides, PPP is the only means available in precarious conditions of practice, including armed conflicts.

## Data Availability

From January 2012, patients of this study were registered in our institution s pelvic trauma registry (authorized by the French National Commission on Data Protection (CNIL) listed under the reference MR-001 No. 1578624vO and in the Traumabase (www.traumabase.eu). Traumabase.eu, created in 2012, is a French network database dedicated to all admissions due to whatever trauma. Traumabase obtained approval from the Advisory Committee for Information Processing in Health Research (CCTIRS, 11.305bis) and from the CNIL (authorization 911461) and meets the requirements of the local and national ethics committees (Comite de Protection des Personnes, Paris VI).

## BIBLIOGRAPHY

1. Filiberto DM, Fox AD. Preperitoneal pelvic packing: Technique and outcomes. Int J Surg. 2016;33(Pt B):222–4.

2. Burlew CC. Preperitoneal pelvic packing for exsanguinating pelvic fractures. Int Orthop. 2017;41(9):1825–9.

3. Ertel W, Keel M, Eid K, Platz A, Trentz O. Control of severe hemorrhage using C-clamp and pelvic packing in multiply injured patients with pelvic ring disruption. J Orthop Trauma. 2001;15(7):468–74.

4. Totterman A, Madsen JE, Skaga NO, Roise O. Extraperitoneal pelvic packing: a salvage procedure to control massive traumatic pelvic hemorrhage. J Trauma. 2007;62(4):843–52.

5. Smith WR, Moore EE, Osborn P, Agudelo JF, Morgan SJ, Parekh AA, et al. Retroperitoneal packing as a resuscitation technique for hemodynamically unstable patients with pelvic fractures: report of two representative cases and a description of technique. J Trauma. 2005;59(6):1510–4.

6. Cotte J, Courjon F, Beaume S, Prunet B, Bordes J, N’Guyen C, et al. Vittel criteria for severe trauma triage: Characteristics of over-triage. Anaesth Crit Care Pain Med. 2016;35(2):87–92.

7. Tile M. Acute Pelvic Fractures: I. Causation and Classification. J Am Acad Orthop Surg. 1996;4(3):143–51.

8. Toulouse E, Masseguin C, Lafont B, McGurk G, Harbonn A J AR, et al. French legal approach to clinical research. Anaesth Crit Care Pain Med. 2018;37(6):607–14.

9. Cothren CC, Osborn PM, Moore EE, Morgan SJ, Johnson JL, Smith WR. Preperitonal pelvic packing for hemodynamically unstable pelvic fractures: a paradigm shift. J Trauma. 2007;62(4):834-9; discussion 9-42.

10. Starr AJ, Griffin DR, Reinert CM, Frawley WH, Walker J, Whitlock SN, et al. Pelvic ring disruptions: prediction of associated injuries, transfusion requirement, pelvic arteriography, complications, and mortality. J Orthop Trauma. 2002;16(8):553–61.

11. White CE, Hsu JR, Holcomb JB. Haemodynamically unstable pelvic fractures. Injury. 2009;40(10):1023–30.

12. Jang JY, Shim H, Jung PY, Kim S, Bae KS. Preperitoneal pelvic packing in patients with hemodynamic instability due to severe pelvic fracture: early experience in a Korean trauma center. Scand J Trauma Resusc Emerg Med. 2016;24:3.

13. Li Q, Dong J, Yang Y, Wang G, Wang Y, Liu P, et al. Retroperitoneal packing or angioembolization for haemorrhage control of pelvic fractures--Quasi-randomized clinical trial of 56 haemodynamically unstable patients with Injury Severity Score >/=33. Injury. 2016;47(2):395–401.

14. Chiara O, di Fratta E, Mariani A, Michaela B, Prestini L, Sammartano F, et al. Efficacy of extra-peritoneal pelvic packing in hemodynamically unstable pelvic fractures, a Propensity Score Analysis. World J Emerg Surg. 2016;11:22.

15. Baque P, Trojani C, Delotte J, Sejor E, Senni-Buratti M, de Baque F, et al. Anatomical consequences of “open-book” pelvic ring disruption: a cadaver experimental study. Surg Radiol Anat. 2005;27(6):487–90.

16. Metz CM, Hak DJ, Goulet JA, Williams D. Pelvic fracture patterns and their corresponding angiographic sources of hemorrhage. Orthop Clin North Am. 2004;35(4):431–7, v.

17. Kim K, Shim H, Jung PY, Kim S, Choi YU, Bae KS, et al. Effectiveness of kaolin-impregnated hemostatic gauze use in preperitoneal pelvic packing for patients with pelvic fractures and hemodynamic instability: A propensity score matching analysis. PLoS One. 2020;15(7):e0236645.

18. Grimm MR, Vrahas MS, Thomas KA. Pressure-volume characteristics of the intact and disrupted pelvic retroperitoneum. J Trauma. 1998;44(3):454–9.

19. Cullinane DC, Schiller HJ, Zielinski MD, Bilaniuk JW, Collier BR, Como J, et al. Eastern Association for the Surgery of Trauma practice management guidelines for hemorrhage in pelvic fracture--update and systematic review. J Trauma. 2011;71(6):1850–68.

20. Spahn DR, Cerny V, Coats TJ, Duranteau J, Fernandez-Mondejar E, Gordini G, et al. Management of bleeding following major trauma: a European guideline. Crit Care. 2007;11(1):R17.

21. Magnone S, Coccolini F, Manfredi R, Piazzalunga D, Agazzi R, Arici C, et al. Management of hemodynamically unstable pelvic trauma: results of the first Italian consensus conference (cooperative guidelines of the Italian Society of Surgery, the Italian Association of Hospital Surgeons, the Multi-specialist Italian Society of Young Surgeons, the Italian Society of Emergency Surgery and Trauma, the Italian Society of Anesthesia, Analgesia, Resuscitation and Intensive Care, the Italian Society of Orthopaedics and Traumatology, the Italian Society of Emergency Medicine, the Italian Society of Medical Radiology -Section of Vascular and Interventional Radiology- and the World Society of Emergency Surgery). World J Emerg Surg. 2014;9(1):18.

22. Suzuki T, Smith WR, Moore EE. Pelvic packing or angiography: competitive or complementary? Injury. 2009;40(4):343–53.

23. Tai DK, Li WH, Lee KY, Cheng M, Lee KB, Tang LF, et al. Retroperitoneal pelvic packing in the management of hemodynamically unstable pelvic fractures: a level I trauma center experience. J Trauma. 2011;71(4):E79–86.

24. Burlew CC, Moore EE, Stahel PF, Geddes AE, Wagenaar AE, Pieracci FM, et al. Preperitoneal pelvic packing reduces mortality in patients with life-threatening hemorrhage due to unstable pelvic fractures. J Trauma Acute Care Surg. 2017;82(2):233–42.

25. Moskowitz EE, Burlew CC, Moore EE, Pieracci FM, Fox CJ, Campion EM, et al. Preperitoneal pelvic packing is effective for hemorrhage control in open pelvic fractures. Am J Surg. 2018;215(4):675–7.

